# The predictive value of arterial stiffness combined with hypertension in the index of atherosclerotic cardiovascular disease

**DOI:** 10.1101/2023.09.14.23295590

**Authors:** Qian Qin, Yang Yang, Jiaoyan Li, Hang Yang, Jingfeng Chen, Yansong Zheng, Suying Ding

## Abstract

**Objective:** This study was conducted to investigate the hypertension and arterial stiffness (AS) in predicting future atherosclerotic cardiovascular disease (ASCVD) risk. **Methods:** We included 6530 participants from Chinese People′s Liberation Army General Hospital. AS was assessed by measuring brachial-ankle pulse wave velocity (baPWV) and participants were stratified into two groups: ASCVD≥10% or ASCVD<10% by a risk threshold of 10%. The Kaplan-Meier survival analysis and Cox proportional risk model were evaluated the risk of ASCVD between participants with ideal vascular function (IVF defined as normal AS with normotension), normotension with AS (NTAS), hypertension with normal baPWV (HTNAS) and hypertension with AS (HTAS). C statistics were used to compare hypertension and AS status in determining ASCVD risk.

**Results:** After a median follow-up of 2.17 years, 672 participants with high risk of ASCVD were identified. Compared to the IVF group, the highest risk of ASCVD was exhibited in the HTAS group (*HR*=2.252, *95%CI*=1.733∽2.927), followed by the NTAS group (*HR*=1.888, *95%CI*=1.583∽2.252) and HTNAS group (*HR*=1.827, *95%CI*=1.144∽2.916). Multiple sensitivity and subgroup analyses yielded similar results. Additionally, compared to the addition of hypertension in the traditional model, the addition of AS elevated the incremental effect on the predicted value of ASCVD (the C statistic was 0.824 vs 0.817, integrated discrimination improvement was 0.80% vs 0.20%, and net reclassification index was 25.00% vs 12.20%).

**Conclusions:** The individuals with AS had a higher risk of ASCVD, and hypertension amplified these associations after adjusting for cardiovascular confounders. Otherwise, AS showed better predictive power than hypertension in determining ASCVD risk.

---

Currently, given that China is challenged by the westernized way of life and population aging, the incidence of metabolic disease is rising steadily, accompany with the increasing in prevalence of cardiovascular disease (CVD). According to the report on Cardiovascular Health and Diseases in 2021, it is estimated that about 330 million patients experienced CVD in China^1^. Furthermore, atherosclerotic cardiovascular disease (ASCVD) accounting for approximately two-thirds of CVD deaths^2^, which has become the leading cause of death worldwide^3^. To put forward effective interventions to deal with the challenges of ASCVD epidemics, it is essential to identify timely the risk features as primary prevention of ASCVD for reducing the prevalence of ASCVD. Majority studies had confirmed that we must take the management of blood pressure as the primary prevention of ASCVD^4, 5^, especially in young adults^6^. Additionally, the arterial stiffness (AS) had been gradually affirmed to be associated with hypertension, type 2 diabetes mellitus and ASCVD^7-9^. Considering that brachial-ankle pulse wave velocity (baPWV) can reflect the function of AS, the baPWV has gradually become an emerging tool for ASCVD risk assessment and stratification in clinical practice^9^. These results imply that both hypertension and AS may predict the risk of ASCVD.

Whereas, study indicated AS, as target organ damage of hypertension, increased the risk of ASCVD^10, 11^, hinting that ASCVD may be attributable to hypertension and its vascular damage. However, the mediating effect analysis indicated that AS appeared to precede the increase in blood pressure^7^. Up to now, whether AS is superior to blood pressure in predicting ASCVD has not been investigated. Therefore, we performed the current study to investigate whether baPWV is a more effective predictor versus blood pressure for assessment the risk of ASCVD based on the Chinese medical examination cohort. This study is aimed to screen high-risk population of ASCVD and put forward constructive strategies to reduce the risk of ASCVD.

## Research Design and Methods

### Study Design and Population

The subjects who underwent physical examination in the department of Health Medicine of Chinese People’s Liberation Army General Hospital from 2009 to 2020 were collected. Questionnaires were filled in, medical history was collected, anthropometric index, biochemical indicators and baPWV were detected. Based on the results of physical examination from 2009 to 2020 as baseline data, and the admission criteria were as follows: subjects underwent arteriosclerosis test with complete data. Exclusion criteria included secondary hypertension, acute cardiovascular and cerebrovascular diseases, malignant arrhythmias, tumors, and chronic kidney disease stage 4-5. All subjects were followed up from 2010 to 2021. This study has been approved by the Ethics Committee of the Chinese People’s Liberation Army General Hospital (S2021-636-01). All participants signed informed consent forms.

### Data collection

After fasting overnight for 8 hours, all participants underwent standardized questionnaires and blood sampling. Their medical history was collected by the unified training of medical personnel. All inspections measured by the same type of inspection equipment and reagents were calibrated uniformly. All test results such as systolic blood pressure (SBP), diastolic blood pressure (DBP), fasting plasma glucose (FPG), glycated hemoglobin (HbA1c), triglycerides (TG), total cholesterol (TC), high density lipoprotein-cholesterol (HDL-C), low density lipoprotein cholesterol (LDL-C), serum uric acid (SUA), Serum creatinine (Scr), homocysteine (Hcy) and hear rates were recorded. In addition, we calculated estimating glomerular filtration rate (eGFR) and mean arterial pressure (MAP).

### BaPWV measurement

BaPWV was measured by the ABI-system 100 Networked AS testing device (Omron Healthcare China Co., LTD.) and performed by specially trained nurses in accordance with the manufacturer’s procedures. In order to keep blood pressure stable, the participants with thin clothes were asked to sit for at least 5 minutes. During the measurement, participants were required to lie down on the examination couch in the supine position and remain quiet. Put cuffs on both arms and ankles, with the lower edge of the upper arm cuffs and ankle cuffs locating in 2-3cm away from the elbow joint and ankle joint, and their air nozzle is aligned with the brachial artery and posterior tibial artery, respectively. Measurements were repeated twice for each at a 5-minute interval, with the second measurement of baPWV were used for analysis. The normal range of baPWV is defined as <1400cm/s, and increased AS is defined as baPWV≥1400cm/s^8, 12, 13^.

### Definition of Group and ASCVD

According to the diagnostic criteria of AS and hypertension, the participants were divided into four groups: ideal vascular function (IVF): baPWV<1400cm/s on both sides with normotension; normotension with elevated arterial stiffness (NTAS): non-hypertension with baPWV≥1400cm/s; hypertension with normal arterial stiffness (HTNAS): hypertension with baPWV<1400cm/s; hypertension with elevated arterial stiffness (HTAS): hypertension with baPWV≥1400cm/s.

The 10-year risk of ASCVD was assessed in accordance with the Chinese Lipid Management Guidelines^14^, which take into consideration factors, such as age, sex, lipid profile, smoking (yes/no), diabetes (yes/no), hypertension (yes/no), and eGFR. After calculating the total score of the above risk factors, the participants were divided into two groups: ASCVD≥10% (high-risk population) and ASCVD<10%.

### Statistical Analysis

The mean ± standard deviation (SD) and the medians with interquartile range were used to described continuous variables. The frequencies and percentages were used to describe categorial variables. One-way analysis of variance (ANOVA), the Kruskal-Wallis test and the chi-square test were used to compare the differences in baseline characteristics of each groups.

Person-years was calculated from baseline to the first assessment of ASCVD (the risk threshold was more than 10%). The incidence rate of high-risk population of ASCVD was calculated by dividing the number of incident cases by the total follow-up duration (person-years). Time-to-event data were evaluated by the Kaplan-Meier method, with the difference between groups compared with the Log-rank test.

Cox proportional hazards regression models were used to compare the risk of ASCVD in each group, and hazard ratios (*HRs*) with 95% confidence intervals (*CI*) were calculated. The IVF group was considered as the reference group in five models which were constructed step-by-step. Model 1 was unadjusted; model 2 was adjusted for age, gender, BMI, WC, heart rate, current smoking and drinking status; model 3 was further adjusted for Hcy, HbA1c, FPG, T-CHO, TG, LDL-C, HDL-C, SUA and Scr. Additionally, in view of baPWV is attributed to distending pressure of the artery, we performed two additional analyses by adjusting MAP (Model 4) and DBP (Model 5), respectively.

Sensitivity analysis were implemented to verify the soundness of the analyses. Firstly, the preliminary analysis was reperformed by redefining risk threshold of ASCVD with more than 10% in 2 consecutive examinations as the dependent variable. Secondly, hypertension was redefined as BP≥130/80 mm Hg. Thirdly, to detect the potential effect of reverse causality, we reperformed the preliminary analysis using a 1-year lag period by excluding incident higher risk populations of ASCVD within the first follow-up. Finally, AS were stratified by a predefined risk threshold of 1400cm/s or 1800cm/s and classified into three groups: baPWV<1400cm/s indicated normal AS, 1400cm/s to 1800cm/s indicated moderate AS, and≥1800cm/s indicated severe AS. Then participants were classified into 6 groups based on the status of hypertension and AS.

Subgroup analysis were performed to verify the ruggedness of the results. According to the SBP<140 mmHg and DBP<90 mmHg as the thresholds, hypertension was categorized into isolated systolic hypertension, isolated diastolic hypertension, and systolic-diastolic hypertension with or without AS to form 6 subgroups, in which IVF group were used as the reference.

Finally, C statistics, integrated discrimination improvement (IDI), net reclassification index (NRI) were used to compare the incremental predictive ability of traditional risk factors and with addition of hypertension, AS or their combined effect.

All analyses were performed using R 4.0.2. A two-tailed test was used and *P*<0.05 was considered statistically significant.

## Results

### Baseline Characteristics

A total of 6530 participants were included in this study. The levels of SBP, DBP, MAP, FBG and TG in IVF, NTAS, HTNAS and HTAS groups were gradually increased (*P*<0.05). In addition, participants in the HTAS group tended to be older, and had the highest age, heart rate, HbA1c, TC, LDL-C and baPWV than other groups. The baseline characteristics across the 4 groups were shown in Table 1.

**Table 1.**
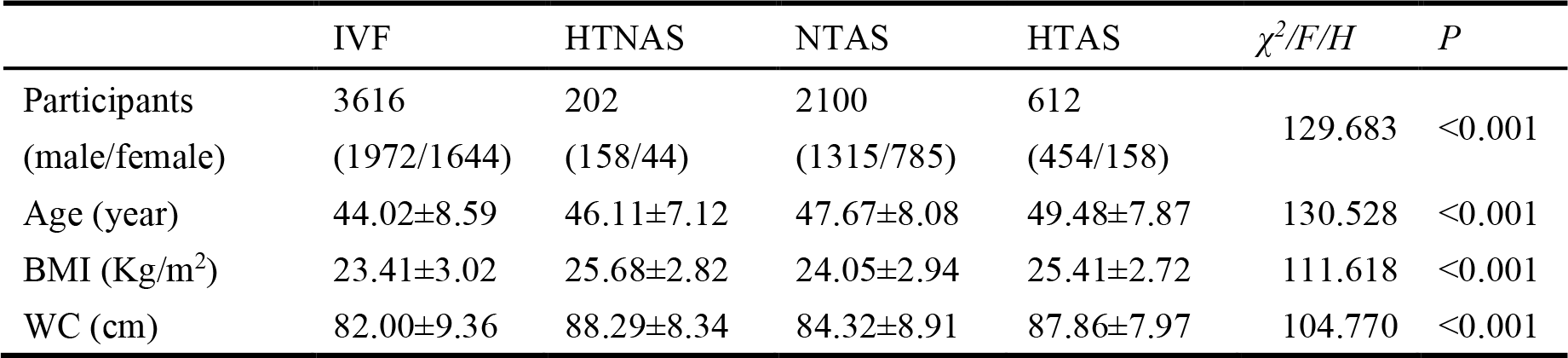
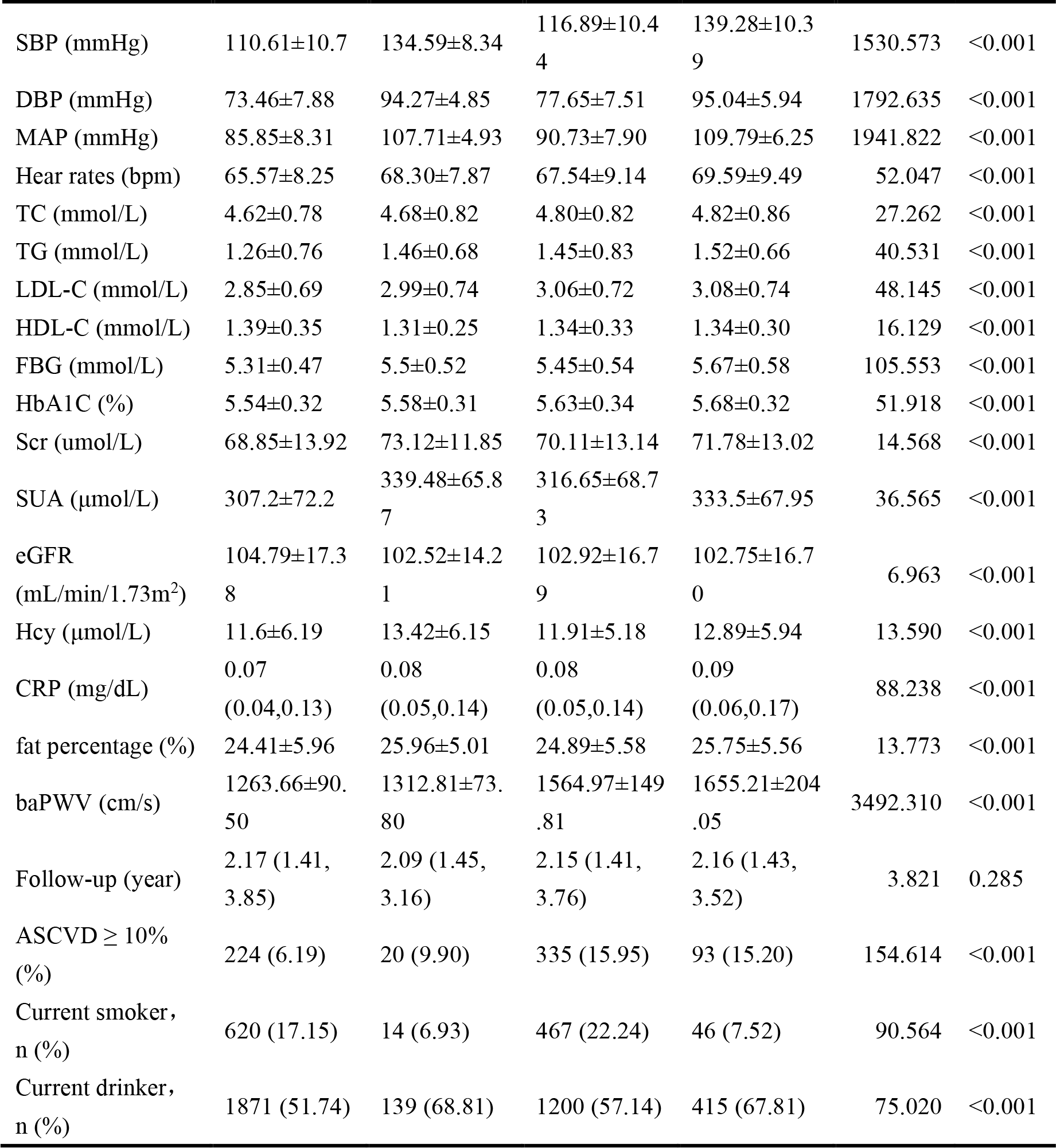
Baseline Characteristics of the Study Population.

### Risk of ASCVD Index in Groups by baPWV and Hypertension Status

The median follow-up of this study was 2.17 years (interquartile range: 1.42, 3.76). By the end of the last follow-up, 672 participants were assessed as high risk for ASCVD. Individuals in the NTAS group suffered from the highest risk of ASCVD than individuals of other groups during the follow-up period (*P*<0.001 for log rank, Figure1), followed by individuals in the HTAS groups. After adjusting for confounding variables in model 3, when compared with IVF group, individuals in the HTAS had the highest risk of ASCVD (*HR*=2.42, *95%CI*=1.93∽3.03), followed by individuals in NTAS group (*HR*=1.888, *95%CI*=1.583∽2.252). The lowest risk of ASCVD was observed in the HTNAS group (*HR*=1.827, *95%CI*=1.144∽2.916). It is worth mentioning that only the individuals in the NTAS groups had the risk of ASCVD after adjusted for MAP or DBP (Table 2), with the HR was 1.692(*95%CI*=1.415∽2.023) and 1.699(*95%CI*=1.420∽2.034), respectively (Table 2).

**Figure1.**
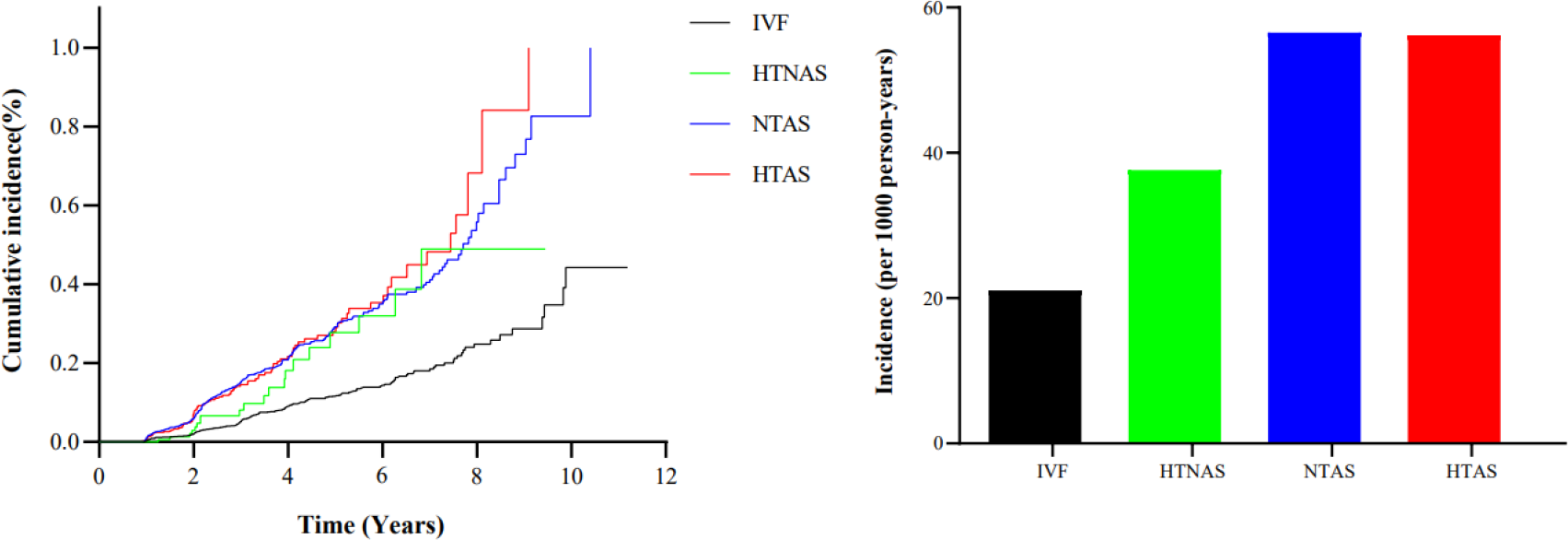
Cumulative incidence of the higher risk of ASCVD by hypertension and AS status. HTAS indicates hypertension with elevated arterial stiffness; HTNAS indicates hypertension with normal arterial stiffness; IVF indicates ideal vascular function; and NTAS indicates normotension with elevated arterial stiffness.

**Table 2.**
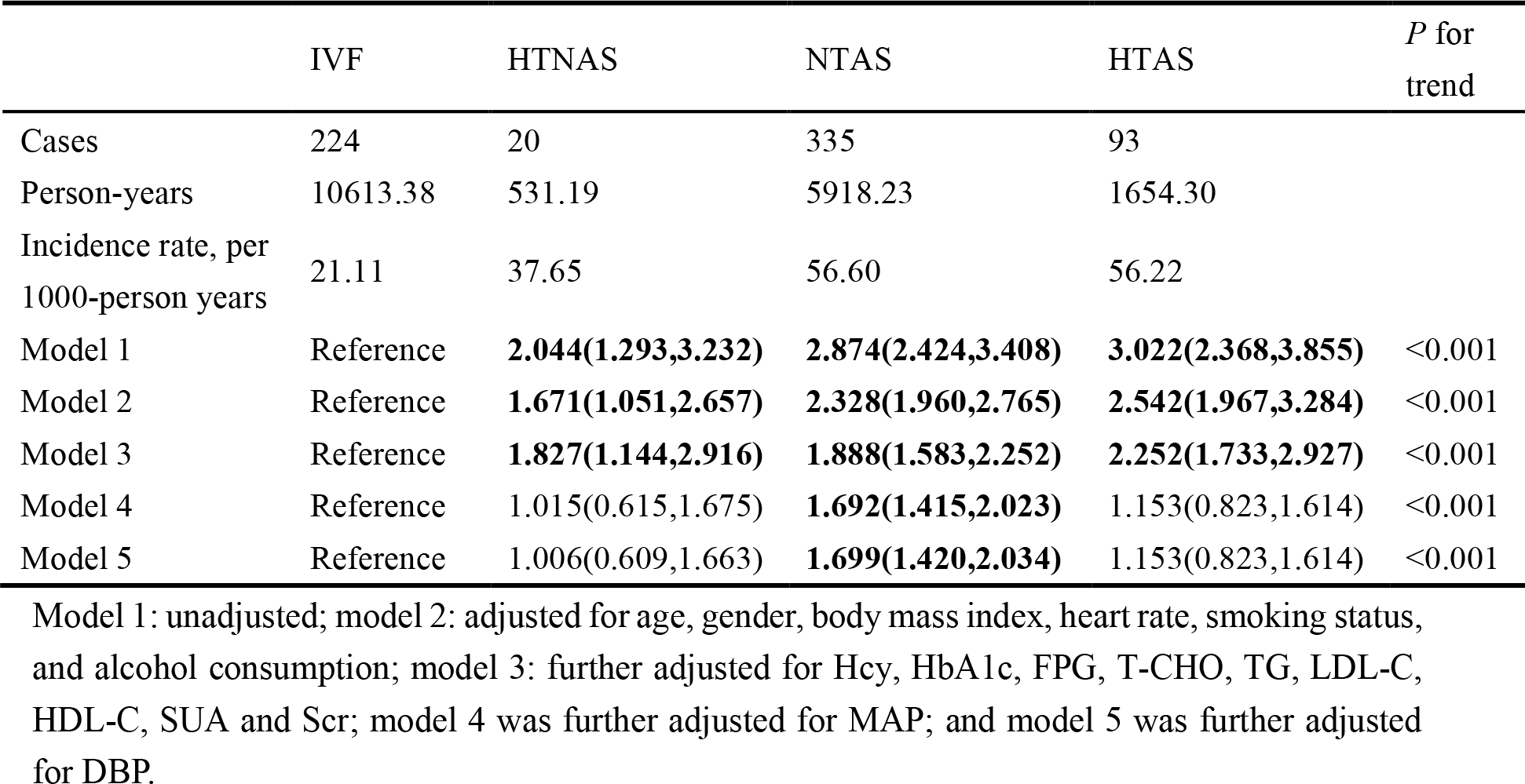
Association of Different Hypertension and Arterial Stiffness Status with Higher Risk of ASCVD.

### Sensitivity analysis

Sensitivity analyses by redefining higher risk individuals of ASCVD in 2 consecutive examinations as the dependent variable (n=111), after adjusting for potential variables, only the participants in the NTAS group emerged higher risk of ASCVD (*HR*=2.316, *95%CI*=1.527∽3.512, Figure2a, Table S1). In addition, sensitivity analysis by reassigning 130/80 mmHg as definition of hypertension, after adjusting for confounding variables, the highest risk of ASCVD was revealed for participants in the HTAS group (*HR*=3.161, *95%CI*=2.508∽3.983), followed by participants in HTNAS and NTAS groups (Figure2b, Table S1). The similar results were also presented in another sensitivity analyses by excluding incident higher risk populations of ASCVD within the first follow-up (n=21, Figure2c). Ultimately, sensitivity analysis by using 1800cm/s as definition of severe AS and 1400cm/s to 1800cm/s as definition of moderate AS, after adjusting for potential variables, the risk of ASCVD was elevated significantly for participants with hypertension and severe AS (*HR*=2.432, *95%CI*= 1.474∽4.012, Figure2d), followed by those with hypertension and medium AS (*HR*=2.208, *95%CI*=1.669∽2.919, Figure2d) and normotension and medium AS (*HR*=1.912, *95%CI*=1.600∽2.286, Figure2d). The lowest risk of incident ASCVD was observed for those with normotension and severe AS (*HR*=1.638, *95%CI*=1.106∽2.426, Figure2d, Table S1).

**Figure 2.**
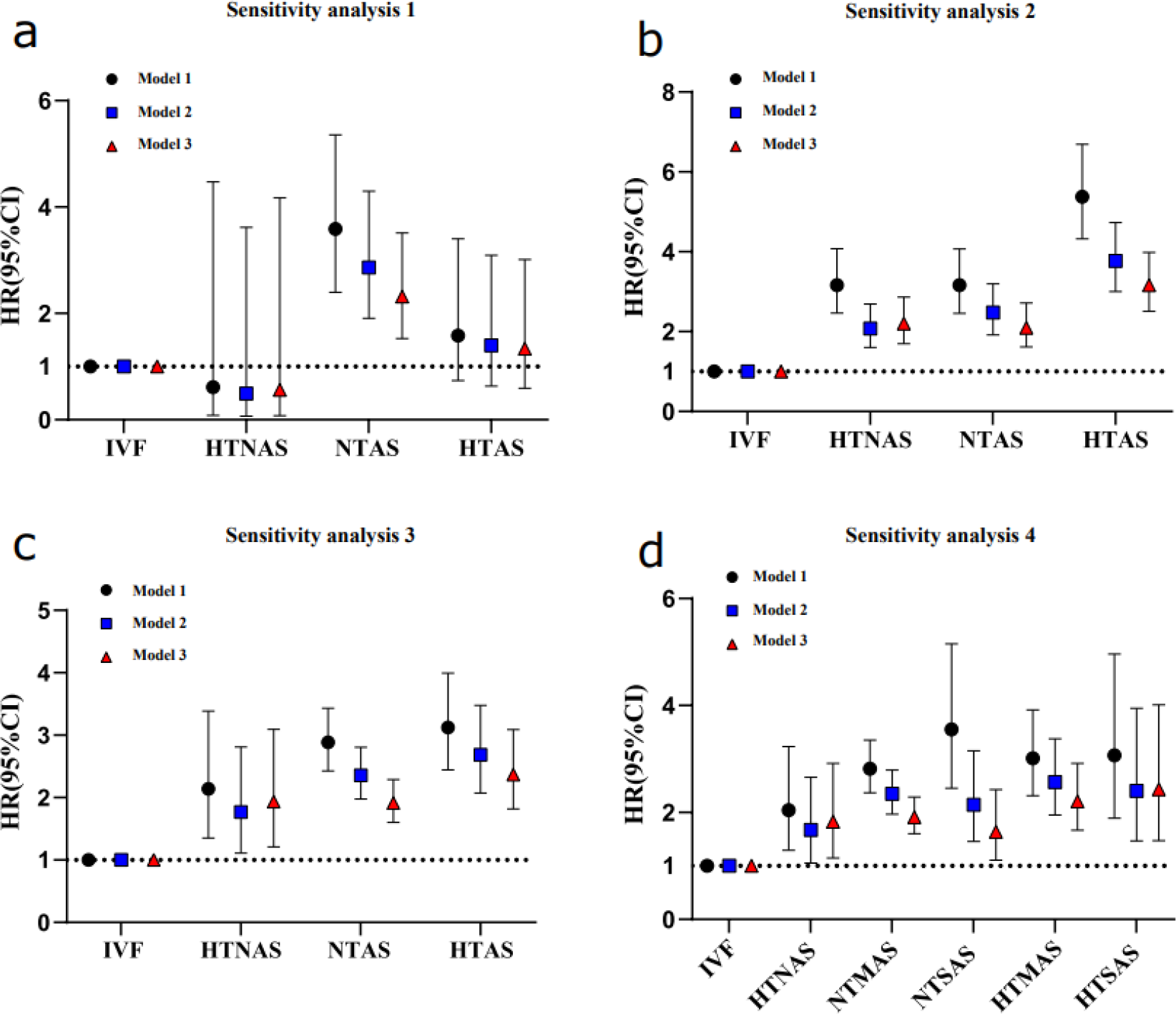
Sensitivity analyses for the association of hypertension and arterial stiffness status with ASCVD. HTSAS indicates hypertension with sever arterial stiffness (baPWV≥1800cm/s); HTMSA indicates hypertension with medium arterial stiffness (1400cm/s<baPWV<1800cm/s); NTSAS indicates normotension with sever arterial stiffness; NTMAS indicates normotension with medium arterial stiffness.

### Subgroup Analysis

The subgroup analysis of subjects with both systolic and diastolic hypertension showed that the higher risk of ASCVD was observed in the HTNAS and HTAS group (Figure 3, Table S2). Unlikely, the subgroup analysis by adopting isolated systolic hypertension and isolated diastolic hypertension in the model, after adjusting for confounding variables, only the subjects in the HTAS group exerted higher risk of ASCVD (Figure 3, Table S2).

**Figure 3.**
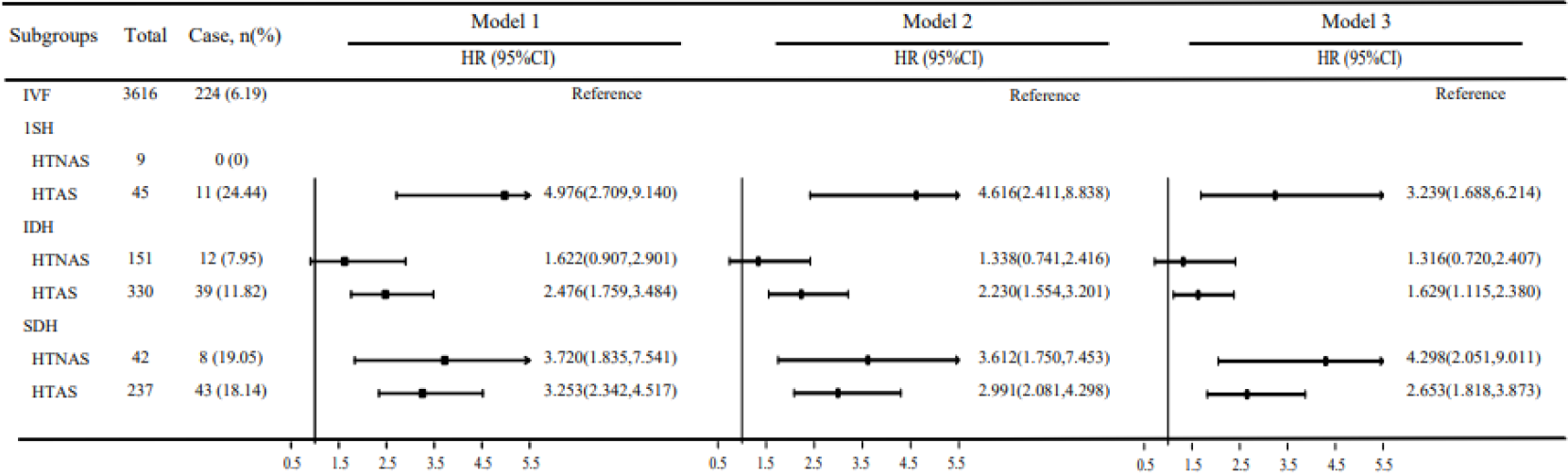
Association of AS status combined with isolated systolic hypertension, isolated diastolic hypertension, and both systolic and diastolic hypertension. In each subgroup analysis, participants with IVF were taken as reference. ISH indicates isolated systolic hypertension; IDH indicates isolated systolic hypertension; SDH indicates both systolic and diastolic hypertension.

### Incremental Predictive Value of AS and Hypertension

The predictive value of hypertension and AS was presented in Table3. The C statistic of the conventional model which was based on the factors of the above model 3 was 0.817(*95%CI*=0.801∽0.833). With the addition of hypertension, the model remained robust, whereas its C statistic did not significantly improve with IDI was 0.114 (*P*>0.05). With the addition of AS, the C statistics were significantly improved from 0.817 to 0.824 (*P*<0.001). The corresponding IDI and NRI were 0.80% (*95%CI*=0.002∽0.015, P<0.001) and 25.00% (*95%CI*=0.183∽0.308, *P*<0.001), respectively. It indicated that the discriminatory power and risk reclassification of AS seemed to be substantial better than that of hypertension. Additionally, with the addition of the combined hypertension and AS, the C statistic was also significantly improved from 0.817 to 0.824, with corresponding IDI was 0.90% (*95%CI*=0.003∽0.018) and NRI was 23.50% (*95%CI*=0.164∽0.300). This result clearly put forward the conclusion that AS performed superior than hypertension in predicting future ASCVD risk.

**Table 3.**
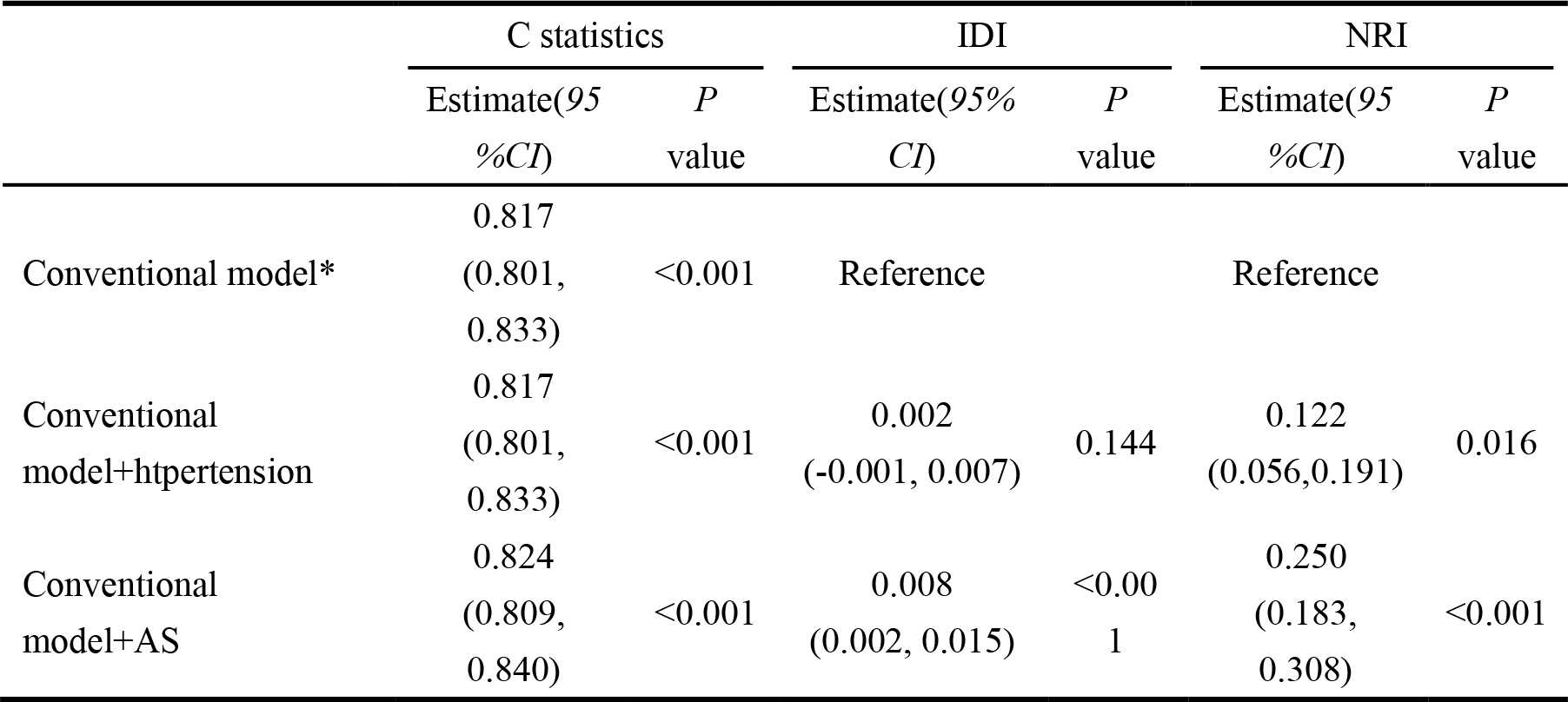
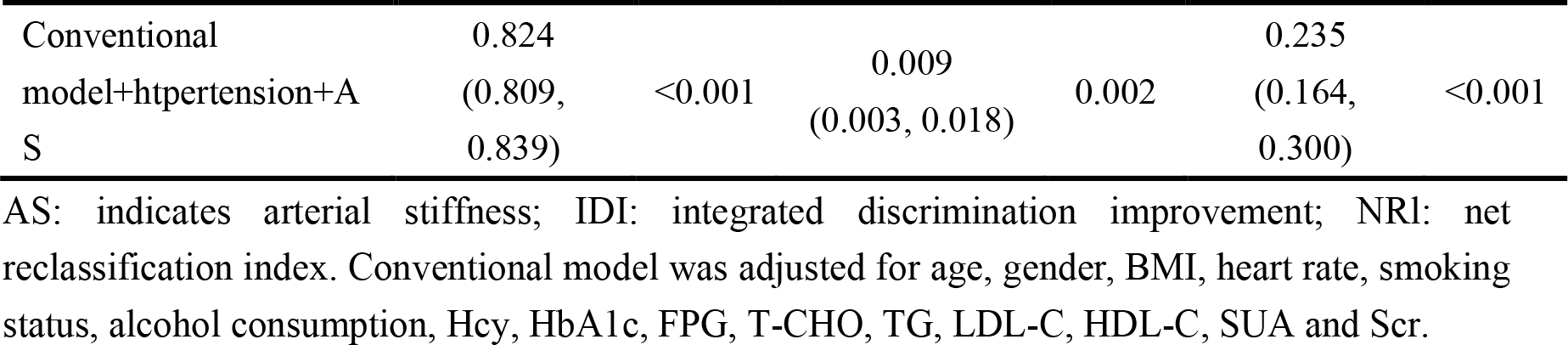
Reclassification and Discrimination Statistics for AS and Hypertension Status.

## Discussion

During the follow-up period, this retrospective cohort study showed participants in the NTAS group underwent the highest risk of ASCVD. Furthermore, when compared with the participants in IVF group, this study demonstrated that participants with AS had a higher risk of ASCVD. Hypertension only amplified these correlations after adjusting for cardiovascular confounders. The results remain steady in sensitivity and subgroup analyses. Ultimately, this study verified AS had a higher incremental ability in determining future ASCVD risk versus hypertension.

Numerous studies, especially the Framingham study^15^, had affirmed that hypertension was a risk factor for ASCVD^4, 5^. In addition, some studies had gradually found that baPWV, as an assessment indicator of AS, can predict all-cause mortality and ASCVD events^16-19^. As early as 2010, a cohort study with 2,642 participants clearly put forward baPWV as an independent predictor of all-cause mortality after 6.5 years of follow-up^17^. Subsequently, another prospective cohort study demonstrated that baPWV was an independent predictor of mortality and cardiovascular morbidity in patients with diabetes^19^. Recently, a large-scale cohort study of meta-analysis provided evidence that adding baPWV was an independent predictor of ASCVD in Japanese individuals without a history of cardiovascular disease^9^. Our findings supported the above hypothesis that AS and hypertension were associated with increasing risk of ASCVD.

Whether ASCVD can be attributed to both hypertension and AS or hypertension and its vessel damage is unknown. Recently, a multicenter observational study had proved that elevated baPWV mediates a positive correlation between aging and blood pressure, indicating AS might precede increased blood pressure^7^. Another clinical trial had discovered that patients with AS (baPWV≥1800cm/s) at baseline were significantly less likely to achieve their target SBP than those without AS, verifying AS consistently preceded SBP^20, 21^. Additionally, a large-scale cohort based on general Chinese population demonstrated that baPWV had a higher predictive value of ASCVD and all-cause mortality versus blood pressure^22^. Those findings again supported our results, which testified that incremental effect of AS is superior to that of hypertension in improving the predictive ability of ASCVD high-risk populations.

It is worth noting that study had indicated that cardiovascular disease risk attributed to AS reduces with age^23^. It is speculated that, on the one hand, there are more cardiovascular risk factors in the elderly, whose synergistic effect may reduce the proportion of AS. On the other hand, elevated baPWV and increased blood pressure were deemed to be related to aging, rather than pathological element. Recently, a study of elderly cohort, with an average age of 71.48 years, presented that elevated baPWV is an independent predictor of all-cause mortality in a community-based elderly population over 10 years of follow-up^18^. However, the exact mechanism of ASCVD caused by AS remains to be elucidated. The changes in arterial elasticity were considered to preceded changes in vascular structure and function. Otherwise, AS leads to the occurrence and progression of arteriole inward remodeling, with increasing resistance and elevated blood pressure, and in turn, central artery stiffness, thus forming a malignant feedback loop^24^.

There are still some limitations in this study. Firstly, this study did not take actual cardiovascular events as the dependent variable. However, there were no substantial change in the sensitivity analysis by excluding individuals assessed the high-risk individuals of ASCVD at the first year and by redefining higher risk individuals of ASCVD in 2 consecutive examinations as the end events. Secondly, the baPWV, rather than carotid-femoral pulse wave velocity (cfPWV) was adopted to evaluate AS. The baPWV had been verified to be closely related to cfPWV^25^, indicating the baPWV was considered as an index of central arterial stiffness. At present, the American Heart Association has included baPWV in the recommended criteria for evaluating AS^26, 27^. Therefore, baPWV is more suitable and ideal screening method for ASCVD in large-scale populations.

In summary, our study based on healthy physical examination cohorts demonstrated that the risk of ASCVD is related to both hypertension and AS. Furthermore, AS was superior to hypertension in determining future cardiovascular risk, which put forward constructive strategies as the primary prevention of ASCVD.

## Data Availability

The data that support the findings of this study are available on request from the corresponding author.The data are not publicly available due to privacy or ethical restrictions.

## Acknowledgments

Acknowledgments: Qian Qin and Yang Yang:Conceptualization, Methodology, Software, Investigation, Formal Analysis, Writing -Original Draft; Jiaoyan Li: Data Curation; Hang Yang: Visualization, Investigation; Jingfeng Chen: Software, Validation; Yansong Zheng and Suying Ding(Corresponding Authors): Conceptualization, Funding Acquisition, Resources, Supervision, Writing -Review & Editing.

## Sources of Funding

Science and Technology Research Project of Henan Province (222102310226); Key Research Projects of Colleges and Universities of Henan Province(21A320035); Henan Province Medical Science and Technology Research Plan Joint Construction Project (LHGJ20210354)

## Disclosures

None.

## Notes

### Competing Interest Statement

The authors have declared no competing interest.

### Funding Statement

This study was funded by the Chinese government(Science and Technology Research Project of Henan Province (222102310226); Key Research Projects of Colleges and Universities of Henan Province(21A320035); Henan Province Medical Science and Technology Research Plan Joint Construction Project (LHGJ20210354))

### Author Declarations

This study has been approved by the Ethics Committee of the Chinese People's Liberation Army General Hospital (S2021-636-01). All participants signed informed consent forms.

